# Remote patient monitoring to interrupt chains of respiratory infections in outpatient care - a case-control study during the 2020/21 infection season

**DOI:** 10.1101/2021.10.08.21264767

**Authors:** Sarah Eichler, Sebastian Carnarius, Edgar Steiger, Dominik von Stillfried

## Abstract

**Aim of the study:** The aim of the study was to investigate satisfaction, saving of time and the possible reduction of patient visits to practices that use Remote Patient Monitoring (RPM) during treatment compared to usual care.

**Methods:** In a case-control study between October 2020 and May 2021, the participating practices were randomized into three groups (two different RPM systems, one control). The doctors were required to enroll patients with acute respiratory infection ≥ 18 years who have a web-enabled device. After a three-month study phase, the doctors were asked to describe the treatment of their patients via online survey. The patients were also questioned. The analysis was carried out descriptively and with group comparisons.

**Results:** 51 practices with 121 patients were included. Overall, the results show a positive assessment of digital care on the patient side. As for the doctors, handling and integration of the systems into consisting practice processes seem to be a challenge. Further, the number of patient visits to the practice was not reduced by using the systems and the doctors did not save time, but the relationship to the patients was intensified.

**Conclusion:** Even if there were no indications for more efficiency by using the RPM systems, the doctors see great potential to intensify the interaction between doctor and patient. In particular, more intensive contact with patients with chronic diseases (e. g. COPD, long-COVID) could be of long-term interest and importance for doctors in outpatient care.

**Trial Registration:** DRKS00023553

## Background

Coping with the COVID-19 pandemic reveals the importance of outpatient care for patients with acute respiratory infections. In countries, where outpatient care could relief and reduce the use of inpatient treatment, a more favorable course of the pandemic was initially observed. [1] Simultaneously, the available intensive care resources were not overused. The outpatient treatment of COVID-19 patients aims to monitor patients in their home environment. Unfortunately, the whole field of research in telemedicine in outpatient care is still too young to provide golden standards, although first promising trials have already been published. [2-6] In addition to the primary function of supporting the quality of care and reducing the risk of infection for patients and doctors, RPM systems offer the opportunity to merge the digitally recorded data in an anonymized or a pseudonymized way to provide real-time insights into the outpatient care of patients with respiratory infections. Many providers have already established themselves on the (German) market over the last years. However, the perception of users might differ from the provider’s assessment of usability. Based on the current data, it is unknown if the systems support reliable data for healthcare. Further, there are not enough research data proving that such systems can be used safely and quickly in the practices and if they might provide more favorable care effects, e.g. a lower admission rate in hospital or a higher satisfaction among the practice team and the patient. Therefore, it was necessary to examine if RPM systems are suitable for reducing patients’ visits to practices and treating patients with respiratory infections in their home environment.

### Aim of the study

The aim of this case-control study was to examine satisfaction, saving of time and the potential reduction of patient visits to practices that use RPM systems compared to usual care.

## Methods

### Study design and endpoints

This case-control study with three groups was carried out from October 2020 to May 2021. Both the doctors and their treated patients were included, if they agreed to participate. Two groups used one of the two RPM systems that were selected in a previous nationwide tender and the third group of doctors treated their patients in usual care without additional digital support. Doctors were required to include patients into the study for a period of three months.

As for the doctors, we specified the following endpoints: effort in patient recruitment and ongoing management effort for monitoring (e.g. due to patient queries), time balance (savings vs. additional work), satisfaction and assessment of the quality of care (e.g. finding critical cases). Concerning the patients’ regards, we wanted to know about their opinions in the quality of treatment (e.g. reduced uncertainty, fears, expenditure of time) and their satisfaction.

### Practices/Doctors

After screening 84 practices/doctors in outpatient care, 51 practices/doctors (general practitioners, internists, ear-nose-throat specialists or pulmonologists treating patients with respiratory infections) were included in the case-control study (Figure 1). After enrollment, practices/doctors were assigned to either to one of the two groups treating patients with a RPM system or the control group using block randomization in the ratio of 1:1:1, based on randomization lists drawn up in advance.

**Figure 1:**
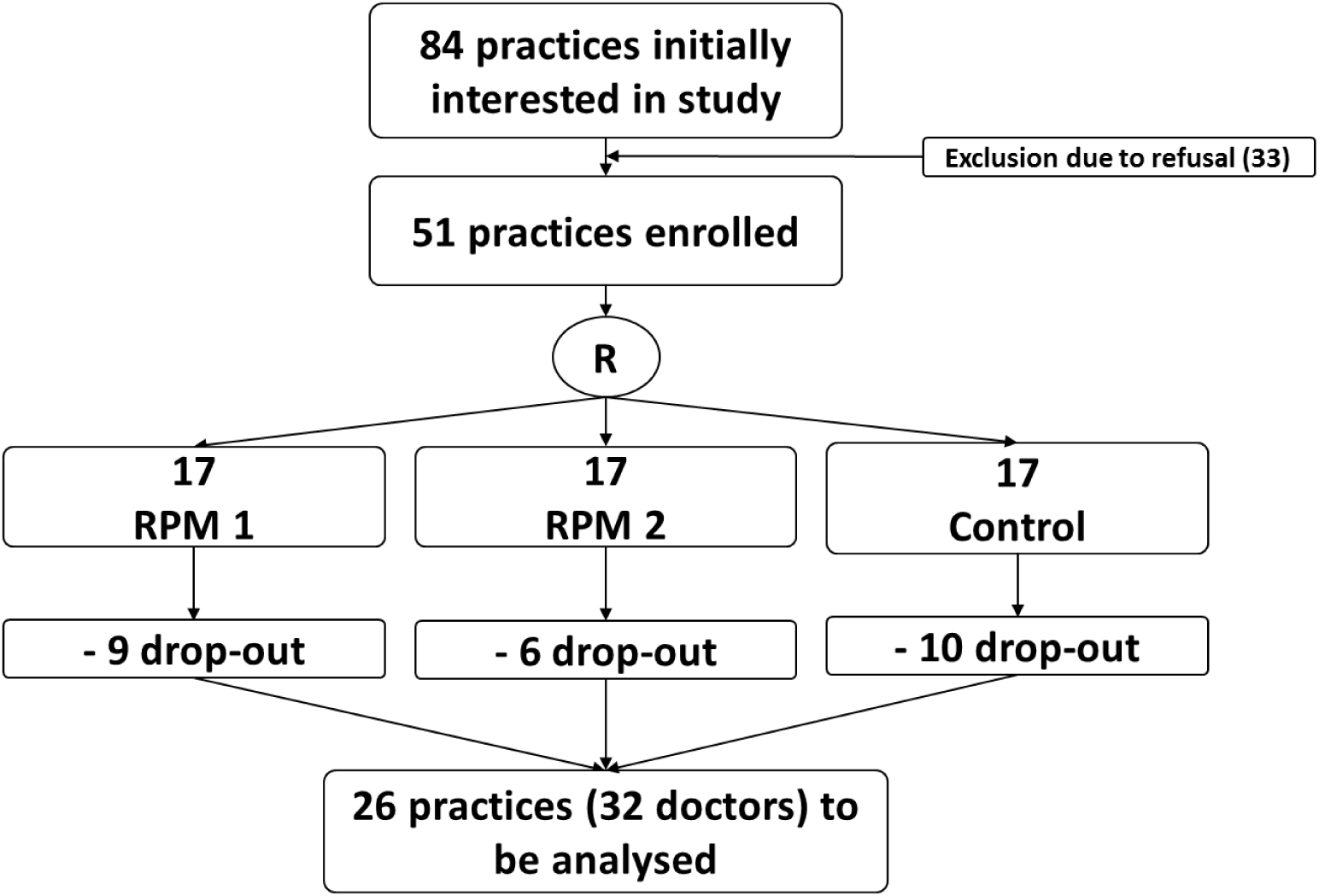
CONSORT flow chart for the inclusion process of practices/doctors.

### Patients

Patients treated in the practices were eligible for inclusion if they were 18 years or older and had an acute respiratory infection. Patients without a web-enabled device were excluded. Insufficient skills of the German language in speech and writing also led to exclusion. Written consent was obtained from all patients. After screening 683 patients, 121 patients could be enrolled. The group they were assigned to was the same as the patients’ investigating doctor (Figure 2).

**Figure 2.**
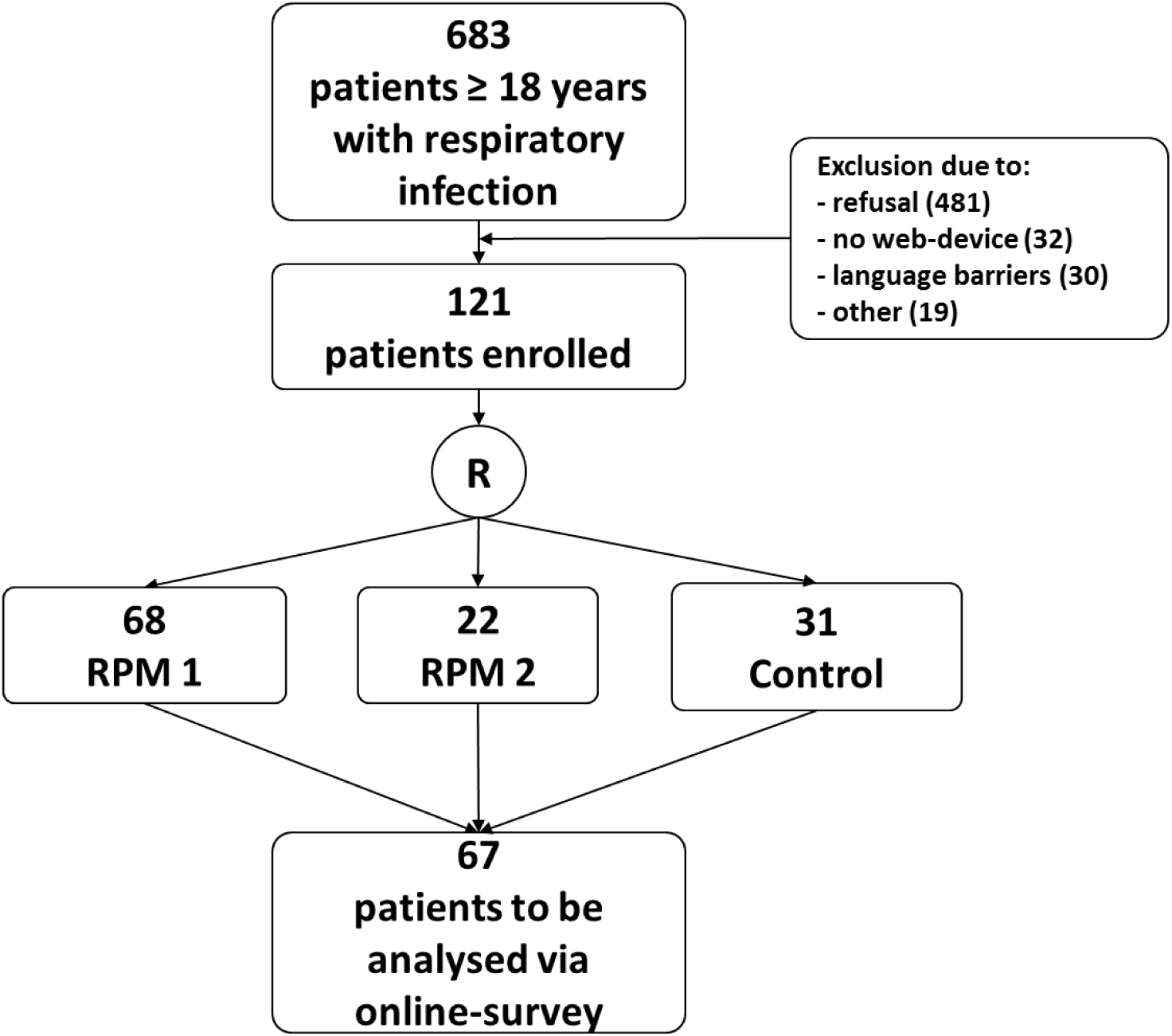
CONSORT flow chart for the inclusion process of patients.

### RPM systems

The two RPM systems used in this study are digital tools that support efficient and safer patient management. They enable the doctor to have higher information density of relevant parameters and information recorded and diagnosed in a shorter time than by patients’ visits in the practice or by phone call. They also enable the continuous recording of parameters, e. g. body temperature, and make the development of the patients’ condition over time visible on the doctors’ dashboard. The RPM could also give the patient feedback that the information transmitted has been viewed by the practice team, so that they can feel safely cared for. The RPM tools therefore should not replace doctor-patient contacts in general, but serve to support the whole treatment. The patients that used one the two RPM systems were instructed in the practices on how to register and to use the systems on their devices. Their treatment lasted as long as the patients’ individual infection. There were no additional interventions or instructions given by the doctor, except the task to constantly add information, e. g. symptoms, requested by the app. After their infection, all patients were asked to fill-out the online survey, whose link they got from their doctor via e-mail.

### Control group

Patients in the control group did not receive any digital support during their treatment. When recovered from their infection, they were also asked to fill-out the online-survey.

### Data collection via online-survey

At the end of the study, the participating doctors were invited to take part in an online survey, which contained 43 questions (19 for the control group) about their characteristics and the endpoints mentioned above. The survey was accessible via a link sent by e-mail from the study site. The doctors were also required to forward the link for the patient survey to their enrolled patients by e-mail. The patient survey comprised 45 questions (26 for the control group) on characteristics and the different endpoints. For many questions, both for doctors and patients, there were seven different possible answers (“totally agree”, “mostly agree”, “tend to agree”, “tend to disagree”, “mostly agree” and “strongly disagree” as well as the field “no answer”). Since the evaluation of these questions showed a differentiated response behavior, the first three possible answers for most questions were categorized as “yes” and the next three as “no” for better presentation; “No information” remained. In this publication, we publish a selection of the most important questions and answers concerning satisfaction and time savings.

### Statistical analyzes

The statistical analyzes were mainly carried out descriptively. For the metric variables, we show mean values and standard deviation and for categorical variables, we demonstrate absolute values and percentages. Different tests for group comparisons were also carried out: analysis of covariance (ANCOVA) for metric variables, χ² tests for categorical ones). The evaluations were performed by using IBM SPSS Statistics 26 and Microsoft Excel 2016.

### Ethics and study registration

A signed informed consent from the patients was required for participation in the study. The patients were informed by their doctor in advance. The study was conducted in accordance with the ethical requirements of the current version of the Declaration of Helsinki. The study procedure and the associated documents were voted positively by the respective ethics committees. Further, the study was registered in the German Clinical Trials Register (DRKS00023553).

## Results

### Doctors’ and patients’ inclusion process and characteristics

84 doctors were interested in participating in the study with their practices. After detailed information on the conditions of the study, 51 doctors/practices agreed to participate. These were randomized into the three groups, so that 17 practices could be assigned to each group. During the three-month intervention phase, 25 practices (49 %) withdrew from the study within the first three weeks, so that the data from 26 practices (51 %) with 32 participating doctors were available for analysis, 13 of them were assigned to RPM group 1, 12 in RPM group 2 and 7 in the control group (Figure 1). 23 (71.9 %) doctors were male and the majority of the doctors (40.6 %) was between 41 and 50 years old. Most doctors (46.9 %) had little experience with clinical studies (1-5 so far) and 27 (84.4 %) were practice owners (Table 1). During the three-month enrollment phase, 683 patients were eligible to participate in the study. The majority (481 patients) refused to participate and 32 patients did not have the technical device needed to participate. 121 patients were enrolled and 67 patients completed the online survey after their infection (22 from RPM group 1, 14 from RPM group 2 and 31 from the control group (Figure 2)). Most of the patients were between 51 and 60 years old and the majority was male (55.2 %). Around two thirds (62.7 %) have not used any health apps and had no chronic diseases (59.7 %). About 9 out of 10 participating patients were non-smokers (85.1 %) and most patients (53.7 %) described the severity of their infection as medium (Table 2).

**Table 1.** Doctor characteristics (n = 32)

|  | **Total cohort**<br>**(n = 32)**<br>**n (%)** | **RPM 1**<br>**(n = 13)**<br>**n (%)** | **RPM 2**<br>**(n = 12)**<br>**n (%)** | **Control**<br>**(n = 7)**<br>**n (%)** | **p-value** |
| --- | --- | --- | --- | --- | --- |
| **Doctors** |  |  |  |  |  |
| Age, years |  |  |  |  | n.a. |
| 18 - 30 | 1 (3.1) | 1 (7.7) | - | - |  |
| 31 - 40 | 3 (9.4) | 2 (15.3) | 1 (8.3) | - |  |
| 41 - 50 | 13 (40.6) | 5 (38.5) | 4 (33.4) | 4 (57.1) |  |
| 51 - 60 | 13 (40.6) | 4 (30.8) | 6 (50.0) | 3 (42.9) |  |
| 61 - 70 | 2 (6.3) | 1 (7.7) | 1 (8.3) | - |  |
| Sex, male | 23 (71.9) | 11 (84.6) | 7 (58.3) | 5 (71.4) | n.a. |
| Practice owner | 27 (84.4) | 11 (84.6) | 9 (75.0) | 7 (100) | n.a. |

Categorical variables are expressed as absolute and relative frequencies with n (%); n.a. = not available due to small number of cases; RPM = Remote Patient Monitoring.

**Table 2.**
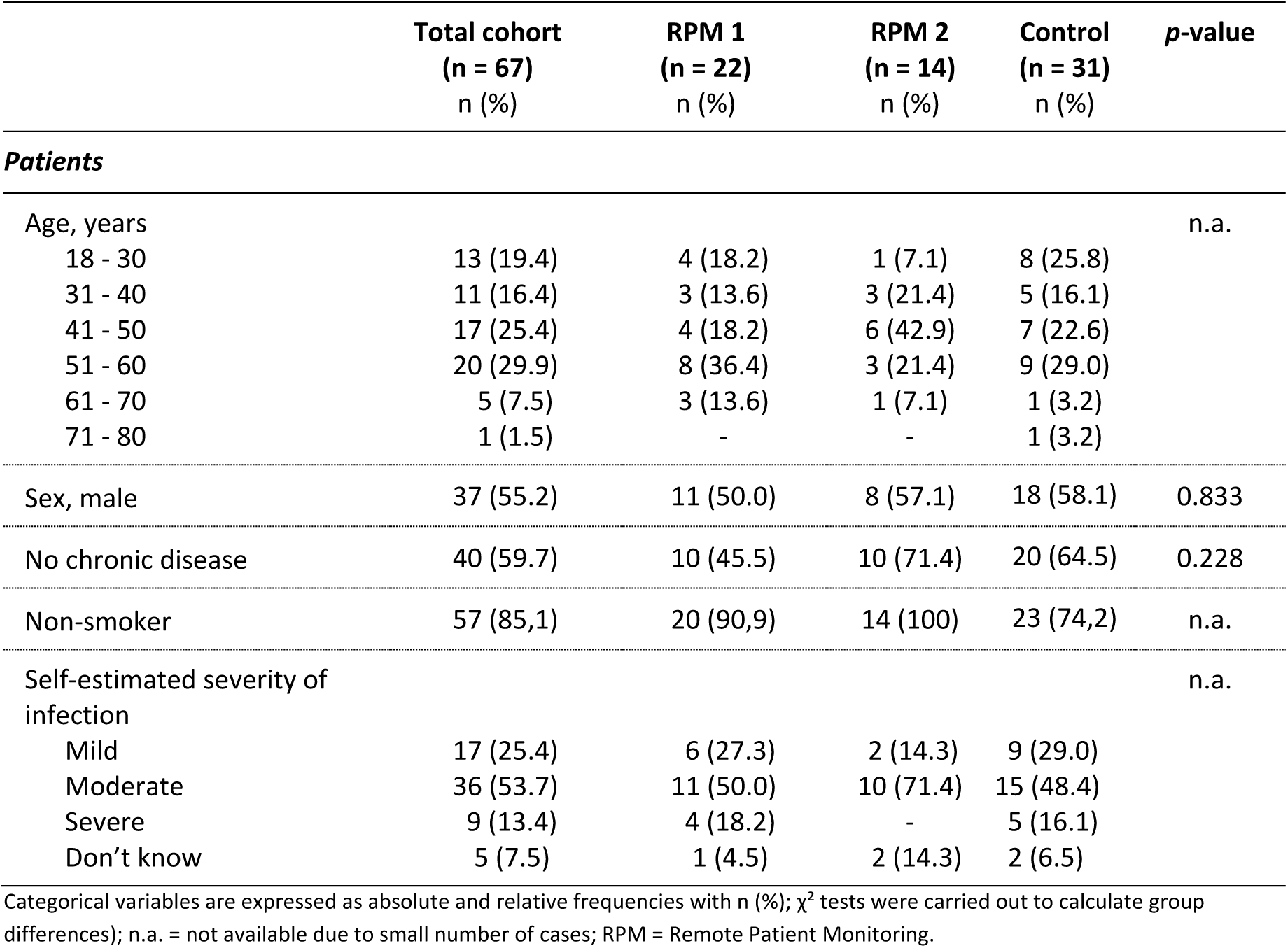
Patient characteristics (n = 67)

|  | **Total cohort**<br>**(n = 67)**<br>**n (%)** | **RPM 1**<br>**(n = 22)**<br>**n (%)** | **RPM 2**<br>**(n = 14)**<br>**n (%)** | **Control**<br>**(n = 31)**<br>**n (%)** | **p-value** |
| --- | --- | --- | --- | --- | --- |
| **Patients** |  |  |  |  |  |
| Age, years |  |  |  |  | n.a. |
| 18 - 30 | 13 (19.4) | 4 (18.2) | 1 (7.1) | 8 (25.8) |  |
| 31 - 40 | 11 (16.4) | 3 (13.6) | 3 (21.4) | 5 (16.1) |  |
| 41 - 50 | 17 (25.4) | 4 (18.2) | 6 (42.9) | 7 (22.6) |  |
| 51 - 60 | 20 (29.9) | 8 (36.4) | 3 (21.4) | 9 (29.0) |  |
| 61 - 70 | 5 (7.5) | 3 (13.6) | 1 (7.1) | 1 (3.2) |  |
| 71 - 80 | 1 (1.5) | - | - | 1 (3.2) |  |
| Sex, male | 37 (55.2) | 11 (50.0) | 8 (57.1) | 18 (58.1) | 0.833 |
| No chronic disease | 40 (59.7) | 10 (45.5) | 10 (71.4) | 20 (64.5) | 0.228 |
| Non-smoker | 57 (85.1) | 20 (90.9) | 14 (100) | 23 (74.2) | n.a. |
| Self-estimated severity of infection |  |  |  |  | n.a. |
| Mild | 17 (25.4) | 6 (27.3) | 2 (14.3) | 9 (29.0) |  |
| Moderate | 36 (53.7) | 11 (50.0) | 10 (71.4) | 15 (48.4) |  |
| Severe | 9 (13.4) | 4 (18.2) | - | 5 (16.1) |  |
| Don't know | 5 (7.5) | 1 (4.5) | 2 (14.3) | 2 (6.5) |  |

Categorical variables are expressed as absolute and relative frequencies with n (%); $\chi^2$ tests were carried out to calculate group differences; n.a. = not available due to small number of cases; RPM = Remote Patient Monitoring.

### Patient visits to the practices

The number of patient visits to the practice could not be reduced by using the systems: patients with a mild course of disease visited the practices 1.5 ± 0.8 times in RPM group 1 vs. 1.3 ± 0.7 times in RPM group 2 vs. 2.3 ± 3.4 times in the control group, p = 0.484 (ANCOVA); patients with a severe course of disease 3.8 ± 1.2 vs. 3.8 ± 2.3 vs. 3.4 ± 1.1 visits, p = 0.902 (ANCOVA), respectively. This is also confirmed by the patients’ answers concerning their number of visits to the practices: 1.7 ± 1.4 vs. 1.7 ± 1.2 vs. 1.6 ± 0.9 vs. 1.7 ± 1.8 visits, p = 0.974 (ANCOVA). Further, according to the statements of the doctors, the systems did not result in less phone calls from worried patients or less need for consultation.

### Saving of time

For most of the doctors, it took a lot of time to instruct the patients on the technology of the RPM systems. In addition, both patient care and practical procedures could not be made more efficient by using the RPM tool. Furthermore, the doctors mainly stated that the RPM systems did not save any time.

### Satisfaction

Both the groups with the RPM systems and the control group stated that they could adequately care for their patients and were satisfied with the care of their patients. The majority of the control group did not want an RPM system for the care of infected patients. The doctors saw RPM tools as fundamentally good support and the doctor-patient relationship was also intensified. The majority of doctors is overall satisfied and would use the system again. The same goes for the majority of the patients, which are overall satisfied, describe their relationship to their doctor as intensified and would use the respective system again.

## Discussion

The RPM study investigated satisfaction, time savings and the possible reduction of patient visits to practices that use RPM systems during the treatment of patients with acute respiratory infections compared to usual care.

All in all, although the effects of the COVID-19 pandemic have accelerated the development of telemedicine in general [7], the infection season 2020/21 represented a special challenge for general practitioners and specialists in the outpatient care. Overall, only a few doctors with their practices were willing to participate in the study, which is certainly also due to the extreme workload of the doctors. Another reason could be, that unclear remuneration or financing of telemedicine is perceived as a barrier [8] and could result in lack of interest in studies investigating telemedicine. Of the few doctors who agreed to participate, 25 practices (49 %) dropped out during the course of the study, all within the first three weeks. It can be assumed that the start, combined with the installation of the systems and the associated training for the providers, but also the documentation effort associated with clinical studies and unavoidable for data protection and ethical reasons (patient information, declaration of consent and patient screening) led to the drop-outs. But also internal practice reasons, general time problems and technical difficulties were given by the doctors. Practices and doctors who have overcome this entry hurdle had no further difficulties in the course of the study.

According to statements by the participating doctors, fewer infection patients were treated in the practices than usual in the infection season examined, but the inclusion rate of the patients is still low at one fifth. There can various reasons for this, e. g. the special situation of the COVID-19 pandemic and the fact that infection patients with less severe courses may not see a need for digital care. Furthermore, patients with severe courses, may have missed outpatient care or were no longer able to digitally document symptoms. It can be assumed that if patients with chronic diseases use these type of RPM systems, it could offer a bigger advantage due to the close relationship with the general practitioner or specialist.

The small number of cases both of the doctors (n = 32) and the patients (n = 67) as well as the different group sizes due to the drop-outs after the start of the study lead to a reduction in the statistical significance. Several tests for group comparisons could not be carried out. Trends observed in this study could turn out to be reliable effects with a larger sample size.

On the patient side, the results show a consistently positive assessment of digital care, even if differences in processing between the two groups that used an RPM system could be relevant in other contexts.

On the medical side, handling and integration of the systems into consisting practice processes in particular still seem to be a challenge. In general, technical maturity is seen as a promotional factor for telemedicine [8]. Furthermore, the use of the RPM systems did not result in a positive time balance in our study. Concerning the positive statements of the control group with regard to the quality and satisfaction of their treatment, the need for digital care options in outpatient care should, if necessary, first be determined separately. Basically, there seems to be a great interest in digital patient care (see also [9]), but in detail there still a problem in the technical implementation. The systems should be able to be integrated into consisting processes as quickly and easily as possible and be able to be used with as little effort as possible by doctors in hectic everyday practice. Otherwise, the inhibition threshold for some doctors may be too high (see also the large number of doctors who dropped out at the beginning of the study).

## Conclusion

Even if there were no indications for more efficiency by using the RPM systems, the doctors see great potential to intensify the interaction between doctor and patient. In particular, more intensive contact with patients with chronic diseases (COPD, long-COVID, etc.) could be of long-term interest and importance for doctors in outpatient care.

## Data Availability

All data produced in the present study are available upon reasonable request to the authors.

